# A high-throughput strategy for COVID-19 testing based on next-generation sequencing

**DOI:** 10.1101/2020.06.12.20129718

**Authors:** Jian Huang, Lan Zhao

## Abstract

COVID-19 testing as sufficient as needed is essential for healthcare workers, patients, and authorities to make informed decisions to confront and eventually defeat severe acute respiratory syndrome coronavirus 2 (SARS-CoV-2). Currently, diagnosis of COVID-19 relies on quantitative reverse-transcription PCR, which is low-throughput, laborious, and often false-negative, making it overwhelmingly challenging to meet testing needs even in industrialized countries. Here we propose a new strategy, which employs a modified loop-mediated isothermal amplification (LAMP) assay, a simple procedure requiring no sophisticated instruments, to index and amplify viral genes from individual specimens, of which the products are readily available for construction of multiplexed libraries for next-generation sequencing. Our strategy would allow precise diagnosis of thousands of specimens in 1-2 days with significantly lower operating expenses. Furthermore, this strategy will make it possible for patients to collect, process, and mail their own samples to facilities for a quick, reliable diagnosis at a population scale.

---

With exponential spread of COVID-19 over every continent except Antarctica, it is fundamentally urgent to ramp up testing capacity for detection of severe acute respiratory syndrome coronavirus 2 (SARS-CoV-2), especially in developing countries which may become new epicenters but have already had their resources stretched to the limit.^1^ Therefore, it is imperative to develop a sensitive, fast, and cost-effective testing strategy to help win this extremely costly war against the coronavirus.

Here we propose a new strategy for high-throughput testing of COVID-19, through combining isothermal amplification and next-generation sequencing. Specifically, we conducted a loop-mediated isothermal amplification (LAMP) assay,^2^ in which viral RNA can be reverse transcribed and amplified at a single temperature (65 °C) in the same vessel, thus greatly reducing the need for sophisticated instruments, as well as minimizing laborious liquid handling that is prone to errors and contaminations. LAMP generates stem-loop DNA of varied sizes from hundreds to thousands of bases (Figure 1), thus they are not directly suitable for downstream applications including sequencing. To overcome this, we introduced recognition sites for PstI and HindIII, both robust, inexpensive restriction enzymes, into the primers of FIP and BIP respectively (Figures 1 and 2), which enabled us to fragment the LAMP products into short (∼150 bp) pieces, thus amenable for adapter ligation and sequencing. We also introduced randomized bases into the primers of FIP, to test whether the addition of nucleotide barcodes impedes LAMP reactions. Our results showed that additions of restriction sites and barcodes in the FIP and BIP primers did not negatively affect amplification (Figure 3), while they offer advantages including barcoding individual samples and fragmenting products in a controlled manner by restriction digestion. Sanger sequencing of LAMP products digested by PstI and HindIII with the FIP or BIP primer confirmed that these short fragments are quite homogeneous target sequences as expected (Figure 3), confirming that LAMP is specific and efficient to amplify traces of viral genes for sequencing. Moreover, if individual samples are indexed by FIP/BIP primers with distinct barcodes (for example, seven-nucleotide barcodes can index over 16,000 specimens theoretically), the opportunity knocks that next-generation sequencing can be applied for a massively parallel detection of COVID-19 in population.

**Figure 1.**
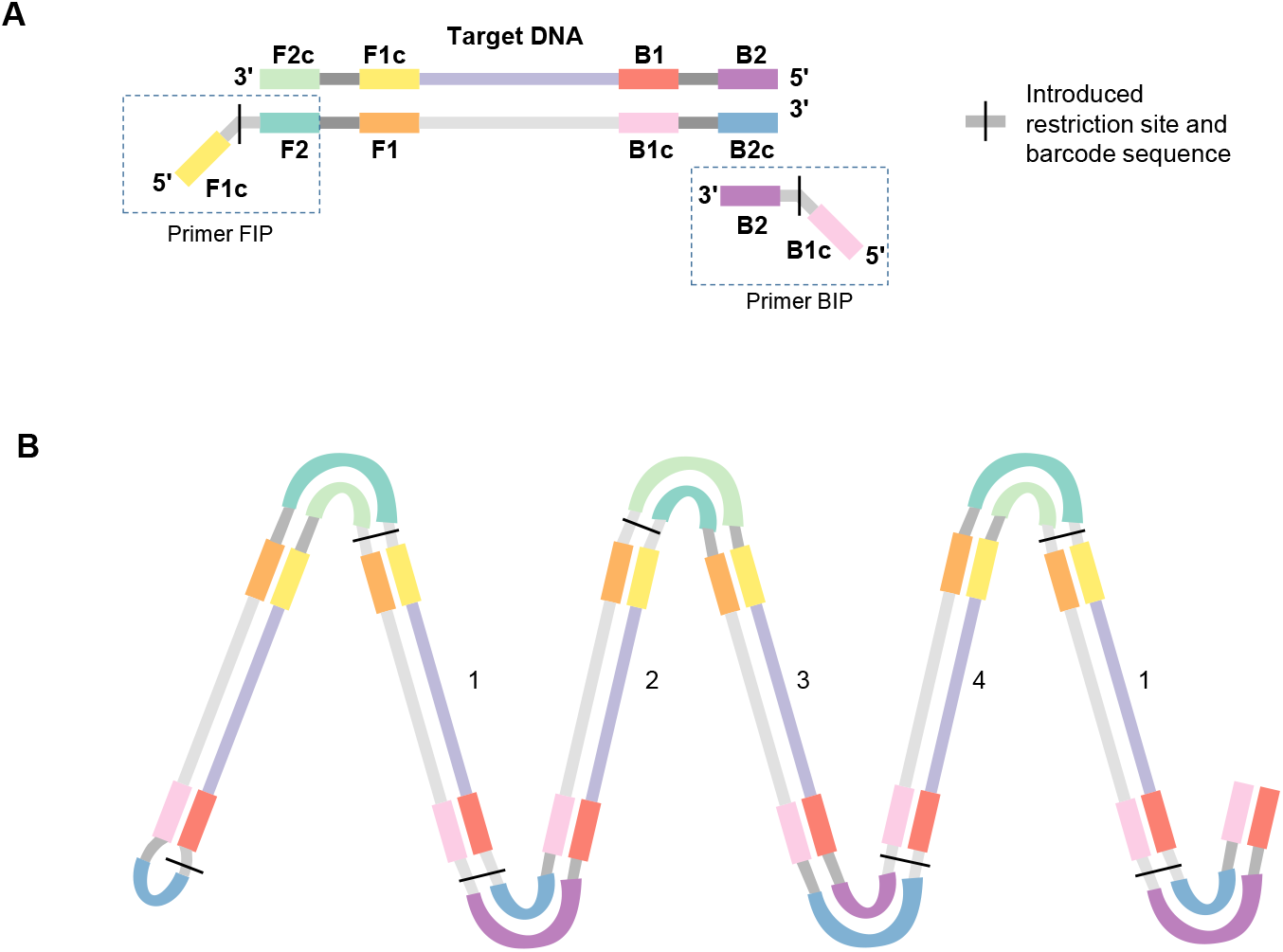
Loop-mediated isothermal amplification (LAMP) can generate individually-barcoded DNA for high-throughput testing based on next-generation sequencing (NGS). (A) Introduction of restriction sites and barcode sequences in the LAMP primer, which does not interfere the reaction but can facilitate population-scale detection of SARS-CoV-2. The structures of the primers are shown as boxed. (B) Schematic of a typical LAMP product. After digested by restriction enzymes, four types of short DNA are released as indicated, which are largely identical and retain the barcodes in two types, suitable for multiplexed NGS library construction.

**Figure 2.**
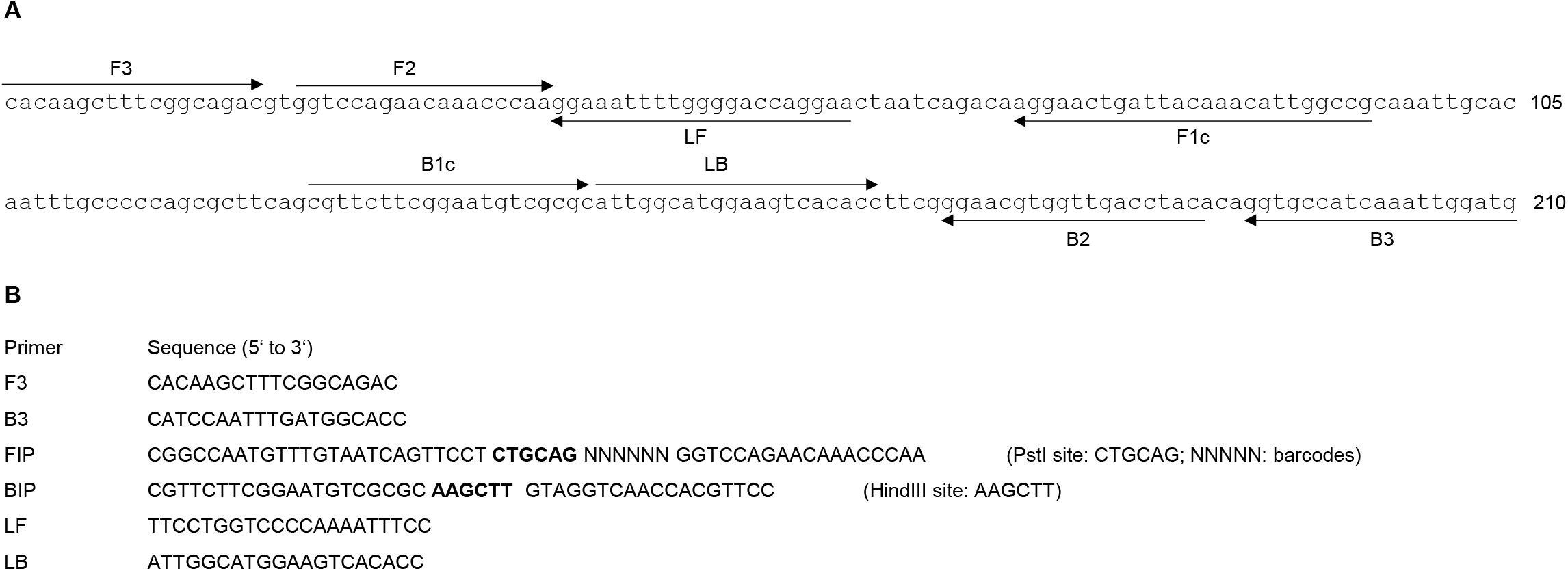
Schematic of LAMP primer design (A) and primer sequences (B) targeting the N gene of SARS-CoV-2. Note the restriction sites in the sequences of FIP and BIP and barcode sequence in FIP.

**Figure 3.**
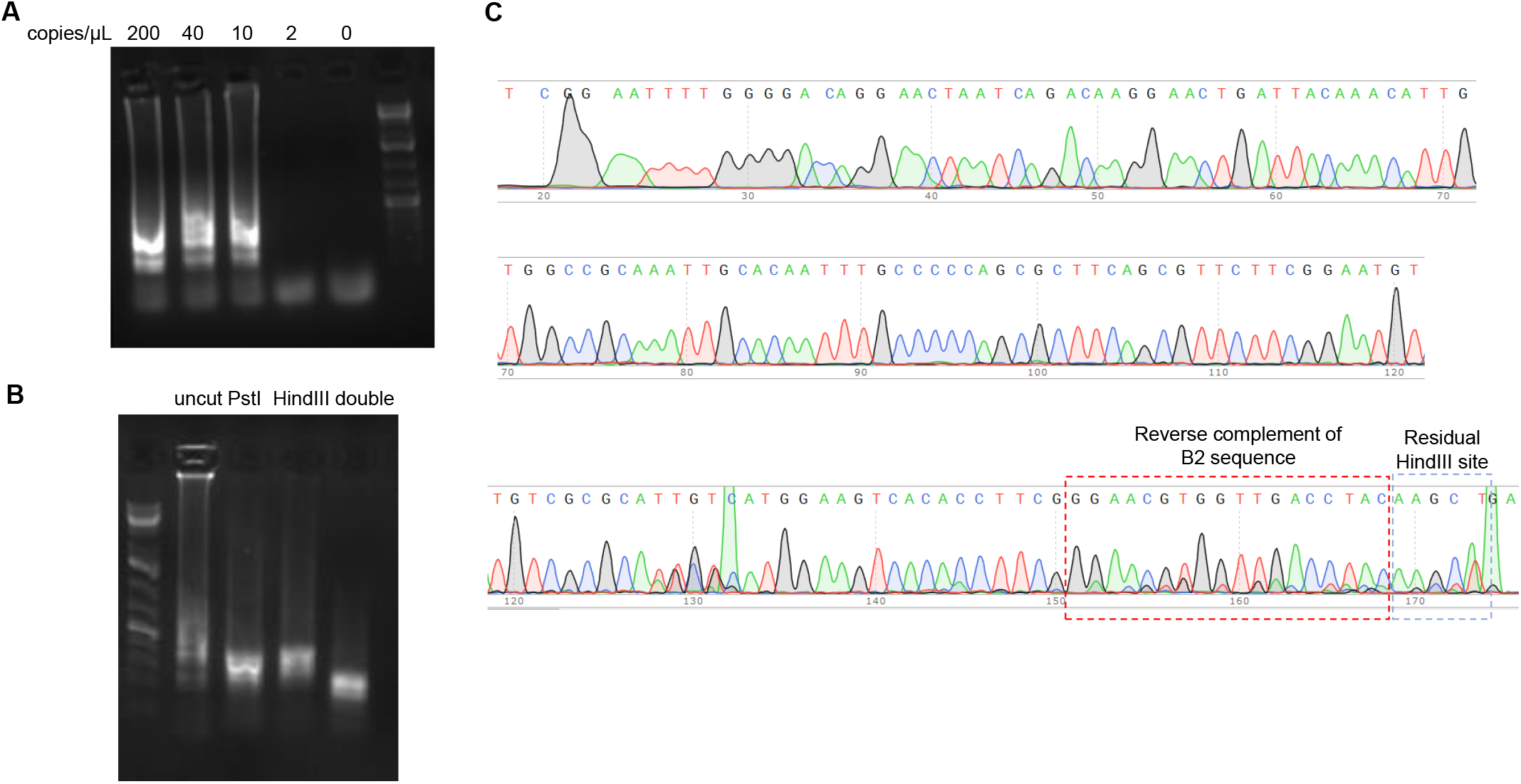
LAMP with barcoded primers is efficient, sensitive and faithful to amplify the targeted sequence. (A) Primers with barcodes and restriction sites amplify the targeted sequence efficiently. (B) Introduction of restriction sites in the primers facilitates fragmentation of LAMP products. Note that digestion by both enzymes generates DNA half the size of those digested by only PstI or HindIII, as expected. (C) Sanger sequencing of the LAMP products digested by PstI and HindIII by the primer FIP confirms that LAMP products are mostly identical to the targeted sequences while retaining the restriction sites.

In summary, we propose that this new testing strategy for SARS-CoV-2 or any other genetic materials can be implemented as follows. Extracted individual RNA (or swab diluent if reaction is further optimized) will be added to the LAMP reaction for reverse transcription and amplification with indexed primers at 65°C for 30 minutes. Then, LAMP products from thousands of specimens will be pooled for fragmentation by restriction digestion, and then ligated to adapters for deep sequencing. Computerized analyses of sequencing results will associate positive specimens with individual patients through the nucleotide barcodes. The complete process will take 1-2 days by 1-2 workers, while fulfilling diagnostic need of thousands of patients with much lower expense compared to current COVID-19 RT-PCR test.

## Discussions

In this study, we introduced restriction sites and barcodes into the LAMP primers, and found that additions of these elements did not negatively affect LAMP efficiency, and the elements were retained in the LAMP products. Sanger sequencing of the LAMP product fragmented by restriction enzymes showed that the LAMP products are quite homogeneous, despite their varying lengths from hundreds to thousands of bases. Thus, our results suggested a potential of LAMP to be used a method to amplify viral genes for NGS. As NGS provides precise sequence information, it eliminates ambiguity caused by nonspecific amplifications in PCR or nonspecific interactions in other diagnostic tests. Moreover, NGS offers a massively parallel capacity to analyze thousands of specimens during a single sequencing run. For high-throughput detection of viral infection in population, the key would be adoption of individual barcode sequences in the LAMP primers. The array of barcoded LAMP reactions can be manufactured in 96-well, 384-well, or any convenient formats. With isothermal amplification and simple handling, hundreds to thousands of specimens can be processed by a worker in a day, which can also be easily automated.

The introduction of restriction sites in the LAMP primers can ensure the fragmentation of LAMP products in a non-randomized manner, and facilitate adapter ligation for NGS by generating sticky ends of the DNA. Nevertheless, barcoded LAMP products can be also randomly fragmented, repaired, and ligated to adapters for NGS using available commercial kits, while possibly possessing lower ligation efficiency and requiring higher sequencing coverage.

In conclusion, our study suggested a great potential of barcoded LAMP reaction to conveniently generate a multiplexed library for NGS, in order to detect COVID-19 in population in a high-throughput manner. It can be even anticipated that patients can collect their own specimens^3^ and process LAMP reactions at home, which are then mailed to facilities to get fast, reliable test results to help them, healthcare workers, and governments to make informed decisions in the fight against SARS-CoV-2 or any other emerging microbes.

## Materials and Methods

### LAMP reactions

LAMP reactions were performed using the WarmStart LAMP Kit (DNA & RNA) (New England Biolabs, E1700S) according to the manufacturer’s instruction. The primer sequences for LAMP reaction were designed using PrimerExplorer V5 (https://primerexplorer.jp) to target the same area in the N gene of SARS-CoV-2 targeted by the CDC-designed N2 RT-PCR assay, with the introduction of PstI or HindIII sites and randomized six bases (Supplemental Figure 1). The positive control for the N gene of SARS-CoV-2 (Catalog # 10006625) were purchased from Integrated DNA Technologies. Specifically, the final concentrations for the LAMP primers were as follows: FIP and BIP, 1.6 µM; F3 and B3, 0.2 µM; LF and LB, 0.4 µM. The reactions were incubated at 65 °C for 30 minutes, and then at 85 °C for 5 minutes to inactivate the enzymes.

### Restriction digestion

After LAMP reactions were completed, 3 µL LAMP products were used for PstI and/or HindIII digestion in a 10 µL volume at 37 °C for 30 minutes. Buffer 3.1 and 0.5 µL of PstI and/or HindIII (New England Biolabs) were used. The digested DNA were analyzed by electrophoresis and gel purified for Sanger sequencing with FIP, BIP, LF, or LB primers. They can be also used to ligate adapters for deep sequencing.

## Data Availability

The data that support the findings of this study are available within the article.

## Acknowledgements

We declare no competing interests. This work was supported by the National Institutes of Health Grants R01AR070222.

## References

1. Global coalition to accelerate COVID-19 clinical research in resource-limited settings. Lancet 2020; 395(10233): 1322–5.

2. Notomi T, Okayama H, Masubuchi H, et al. Loop-mediated isothermal amplification of DNA. Nucleic Acids Res 2000; 28(12): E63.

3. Tu YP, Jennings R, Hart B, et al. Swabs Collected by Patients or Health Care Workers for SARS-CoV-2 Testing. The New England journal of medicine 2020.

